# Quantitative T_2_ Brain Mapping with Simultaneous RF Estimation Using Dual Interleaved Steady States at 7T MRI

**DOI:** 10.64898/2026.03.27.26349590

**Authors:** Dana Yacobi, Rita Schmidt

**Author notes:** **Corresponding author:** Dr. Rita Schmidt.

## Abstract

**Objective:** Quantitative T₂ mapping plays a critical role in brain imaging for assessing a range of neurological conditions, including neurodegenerative diseases, demyelinating disorders, and cerebrovascular pathologies. Despite its diagnostic potential, implementing quantitative T₂ mapping at ultra-high magnetic field strengths (≥7T) poses significant challenges. These include elevated specific absorption rate (SAR) and radiofrequency (RF) field inhomogeneities, which can lead to prolonged scan durations and inaccuracies in quantification.

**Materials and Methods:** Phase-based gradient-recalled echo (GRE) techniques have recently emerged as promising rapid acquisition with enhanced sensitivity to T₂-related contrast. In this study, we introduce TWISTARE (TWo Interleaved Steady-states for T₂ and RF Estimation), a novel dual steady-state 3D-GRE approach that employs interleaved flip angles and small RF phase increments to jointly estimate T₂ and B_1_ maps. By combining two dual-steady-state scans, TWISTARE enables fast, whole-brain quantitative T₂ mapping while reducing scan time and mitigating B_1_-related bias at ultra-high field.

**Results:** Validation experiments included Bloch simulations, phantom studies and in-vivo imaging. The results demonstrated high precision in phantom experiments, achieving up to a two-fold reduction in acquisition time and achieved precision comparable to the gold-standard method in vivo within a similar scan duration.

**Discussion:** TWISTARE establishes a fast steady-state framework for quantitative neuroimaging at ultrahigh field, offering potential benefits for both clinical and research applications, especially in longitudinal and dynamic studies of brain tissue.

## Introduction

Quantitative magnetic resonance imaging (qMRI) provides a powerful neuroimaging framework for the non-invasive, in vivo assessment of brain tissue microstructure and biochemical properties. Among the principal qMRI parameters, the T_2_ relaxation time offers critical insights into tissue composition and has been widely applied in the diagnosis and monitoring of neurological conditions, including neurodegenerative diseases, stroke, multiple sclerosis, and iron accumulation^1–7^. T₂ mapping serves as a quantitative fingerprint across clinical and research settings, supporting investigations of physiological changes, aging, and pathology, and enabling robust longitudinal and multi-site studies^8–13^.

Traditional spin-echo (SE)-based T₂ mapping^14^ remains the gold standard due to its robustness and accuracy. However, the need to acquire multiple spin-echo images at different echo times (TEs) in separate scans leads to prolonged scan durations and increased RF energy deposition. Multi-echo spin-echo (ME-SE)^15^ sequences mitigate scan time constraints by acquiring multiple echoes per excitation, but this comes at the cost of substantially increased RF power deposition. Moreover, ME-SE acquisitions introduce stimulated echoes and complex coherence pathways contributions that may bias T₂ estimates. Recent studies have addressed these effects using dictionary-based approaches and extended phase graph (EPG) modeling to improve T₂ estimation^16–18^. Accelerated strategies have also combined spin-echo with single- or multi-shot echo-planar imaging (EPI)^19–24^, enabling faster acquisition and multi-contrast outputs.

Alternative approaches include fast steady state pulse sequences such as balanced steady-state free precession (bSSFP)^25,26^, DESPOT2^27,28^, and Triple-Echo Steady-State (TESS)^29,30^. Novel approaches, including variable flip angle techniques^31,32^ and T₂-preparation pulse methods^33,34^, have been developed to generate robust T_1_ and T_2_ maps. An emerging and powerful framework is Magnetic Resonance Fingerprinting (MRF)^35,36^, which employs time-varying acquisition schemes and dictionary-based matching to simultaneously estimate T₁, T₂, and B₀. Although MRF demonstrates substantial potential, it remains computationally demanding and may require prolonged acquisition times for 3D imaging, particularly at high spatial resolution.

While most methods rely primarily on the magnitude of the MRI signal, several approaches have also explored the use of phase information^37^. Recently, phase-based gradient-echo (GRE) techniques have been developed using short repetition time (TR) and small RF phase increments during the RF pulse train to preserve coherent transverse magnetization, enabling rapid T_2_-weighted phase imaging^38–40^. Initially assuming a spatially uniform and a priori known flip angle, these methods were subsequently extended to support simultaneous B₁ mapping^41^. More recently, magnitude information has been incorporated into the model fitting framework, allowing simultaneous estimation of T₁, T₂, B₁, and proton density (PD)^42^.

The advent of ultrahigh-field MRI (≥7 Tesla) has significantly advanced brain imaging by providing increased signal-to-noise ratio (SNR) and contrast-to-noise ratio (CNR), thereby enabling the detection of subtle tissue alterations and small lesions that may be imperceptible at lower field strengths^43^. However, these advantages are accompanied by significant technical challenges, particularly radiofrequency (RF) field (B_1_) inhomogeneity and elevated specific absorption rate (SAR), both of which complicate accurate T₂ quantification at 7T. A major limitation at ultrahigh field is RF field inhomogeneity, which arises as the RF wavelength approaches the dimensions of the human head^44^. This results in spatially non-uniform flip angle distributions across the brain^45–47^, introducing location-dependent errors in relaxation time measurements and thus compromising the accuracy of quantitative maps. In addition, increased SAR at 7T imposes stringent constraints on conventional T₂ mapping sequences^47^, particularly those requiring multiple refocusing pulses. To address these challenges, recent 7T studies have explored parallel transmit (pTx) strategies for B_1_ homogenization^48,49^ and incorporation of a priori B_1_ map into T_2_ estimation^50^.

In this study, we propose a novel dual steady-state technique termed TWo Interleaved Steady-states for T₂ and RF Estimation (TWISTARE). The proposed method extends phase-based GRE approaches by employing interleaved flip angles within a 3D GRE sequence and incorporating both magnitude and phase information into a unified estimation framework. TWISTARE enables simultaneous T₂ and B₁ mapping at 7T using only two 3D acquisitions. Whereas the previous approach^41^ required three to four separate scans to remove B₀ contributions and reconstruct T₂ and B₁ maps, reducing the acquisition to two scans improves efficiency and enhances robustness to inter-scan variations, such as subject motion and B₀ fluctuations. We present the theoretical framework of TWISTARE and validate its performance through numerical simulations, phantom experiments and human brain imaging at 7T MRI.

## Methods

### TWISTARE Pulse Sequence Design

TWISTARE is a two-scan dual steady-state acquisition implemented using a gradient-spoiled 3D GRE sequence with a short TR (TR = 10 ms) and small RF phase increments applied throughout the RF pulse train. Unlike conventional RF spoiling, TWISTARE employs small phase increments (0.5° and 3°), which preserve T_2_-sensitive transverse coherence pathways and enable joint estimation of T_2_ and B_1_. The sequence employs interleaved flip angles, such that odd excitations are applied with flip angle α_odd_ and even excitations with flip angle α_even_. This alternating excitation scheme establishes two distinct steady states, generating corresponding odd and even magnitude and phase images. The complex signal for each steady-state and scan can be expressed as

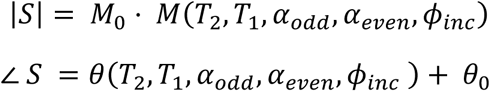

where |S| and ∠S denote the signal magnitude and phase, respectively; *M*_0_ represents the proton density; and *θ*_0_ corresponds to the B_0_-dependent phase contribution. From the resulting signal states, magnitude ratio (MR) and phase difference (*Δθ*) are computed:

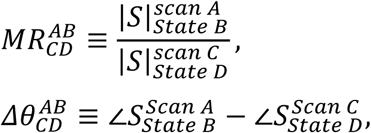

where A and C denote scan identifiers and B and D represent steady-state identifiers. Previous phase-based approaches used two separate acquisitions with positive and negative phase increments to eliminate *θ*_0_. The dual steady-state design enables intrinsic *B*_0_ cancellation through intra-scan phase subtraction while removing *M*_0_ contributions via magnitude normalization.

The selected scan configuration and processing workflow are illustrated in Fig. 1.

**Figure 1:**
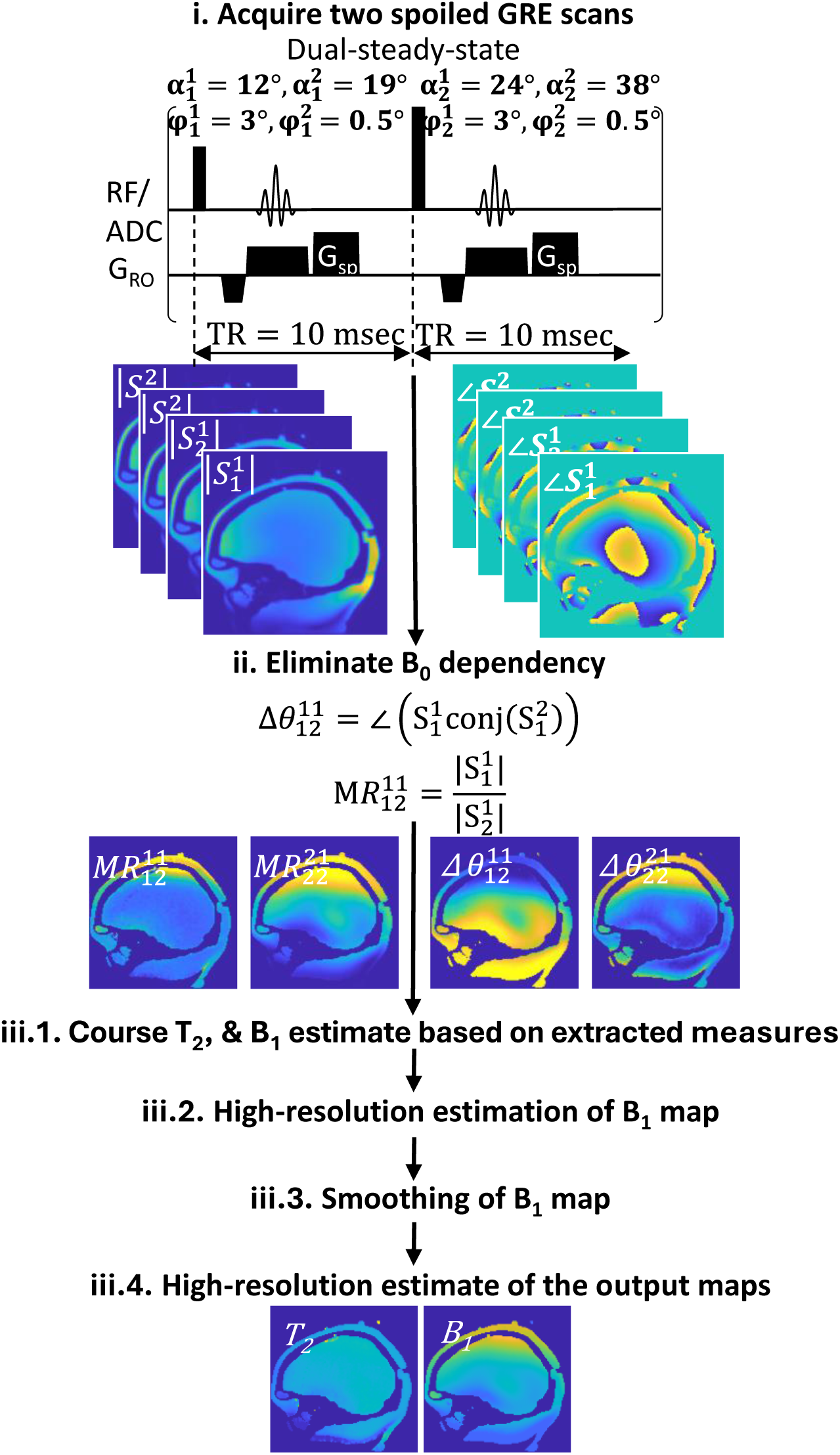
Flowchart illustrating the TWISTARE method, including scan diagram, acquisition parameters, acquired datasets, estimation pipeline, and representative input and output maps.

### Bloch Simulation and dictionary generation

To characterize the dual-steady-state signal behavior, 1D single-voxel Bloch simulations were performed. Steady-state was defined as a relative signal change of less than 0.1% between fifty consecutive excitations. The number of initial repetitions (“dummy scans”) required to reach steady-state was determined accordingly, with a maximum limit of 10,000 repetitions. Once steady state was reached, a single acquisition was simulated.

This procedure was repeated over a multidimensional parameter grid spanning flip angles from 0° to 30° (1° increments), T₂ values from 0 to 200 ms (5 ms increments) and from 0.5 to 4 s (0.5 s increments), and T₁ values from 0.5 to 3 s (0.1 s increments) and from 3 to 6 s (1 s increments). Simulations were performed for multiple RF phase increments (φ_inc_) and amplitude ratios between the two interleaved flip angles. Figure 2 illustrates the phase dependence of odd and even steady states, compared to a single steady state, as function of T₂ and flip angle, highlighting the distinct signal fingerprints that enable parameter estimation.

**Figure 2:**
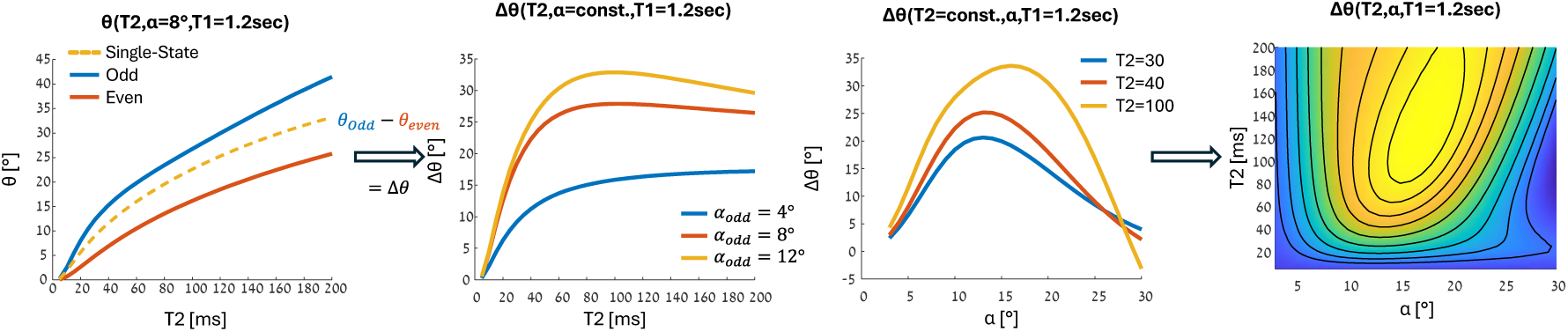
Simulated signal phase in single- and dual-steady-state configurations with φ_inc_=0.5°. From left to right: (1) 1D θ(T_2_) for single-state (α=8°) and a dual steady-state (α_odd_=8°, α_even_=16°); (2) phase difference between states Δθ(T_2_) for α_odd_=4°,8° and 12° (with α_even_=2· α_odd_); (3) Δθ(α_odd_) for T_2_=30ms, 40ms and 100ms (α_even_=2· α_odd_) and (4) 2D map of Δθ(T_2_,α_odd_) (α_even_=2· α_odd_). All simulations were performed with T_1_=1.2 s.

For the selected phase increments and flip angle ratios, the final dictionaries were interpolated with resolutions of 0.1 ms in T₂, 0.1° in flip angle (α), and 100 ms in T₁, enabling precise voxel-wise matching during TWISTARE-based parameter estimation.

### Estimation process

The generated dictionaries were used to estimate T₂ and α at each voxel, based on six phase difference measurements 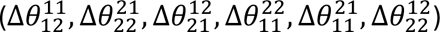 and six magnitude ratio measurements 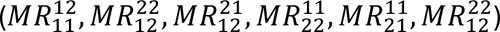. Because these observables differ both in numerical scale and in their sensitivity to T_2_ and the flip angle parameters, a weighting scheme was introduced to balance their influence. Weights were derived from Bloch simulations incorporating additive Gaussian noise to reflect realistic SNR conditions (see Supplementary Information S1).

The parameter estimation followed a four-step procedure: i) initial estimation of α, T₂, and T₁ using coarse (nearest-neighbor) dictionary matching; ii) refined estimation of α using high-resolution dictionaries in a smaller window centered on the previously estimated value; iii) spatial smoothing of α using a 3D Gaussian filter; and iv) final re-estimation of T₂ using the high-resolution dictionary with α fixed to the smoothed values from the previous step. Note that, while the chosen scan parameters do not provide sensitivity to T₁, T_1_ was nevertheless included in the dictionary. Figure 1 shows examples of phase-difference and magnitude-ratio maps acquired in a phantom, from which T_2_ and flip angles were estimated. Note that, due to phase convergence and reduced sensitivity at higher T₂ values, the T₂ estimation range was limited to 200 ms.

In TWISTARE, T_2_ estimation can be performed using either the positive or negative phase increment configuration independently. Both datasets were therefore acquired. T_2_ maps were reconstructed separately from each configuration and additionally from a combined dataset. For the combined dataset, the T_2_ estimate was calculated as the average of the estimates obtained from the positive-ϕ_inc_ and negative-ϕ_inc_ acquisitions.

To mitigate inter-scan B_0_ variations (ΔB_0_), a dual-echo acquisition was implemented. Phase offsets, estimated as Δθ_0_=2π ΔB_0_·TE_echo1_, were corrected, thereby improving robustness to physiological phase fluctuations due to motion and respiration. This correction was applied in vivo analysis.

### Scan Parameter Optimization

To optimize the performance of the TWISTARE method, extensive Bloch simulations were conducted to identify the combination of flip angle ratios and RF phase increments that maximized the sensitivity and accuracy for T₂ and B_1_ mapping. The simulations incorporated a range of tissue properties representative of human brain relaxation times to ensure robustness across different tissue types.

Simulations were performed over a broad range of T₂ values (3 to 200 ms) and flip angles (1° to 30°). Each condition was repeated 1000 times with Gaussian-distributed noise added to the complex signal to assess bias and precision under realistic noise conditions. The noise level was set to match the SNR assumptions used in the weighting procedure, corresponding to an SNR of 100 in a representative central white matter voxel (T₁ = 1.2 s, T₂ = 38 ms), with α = 10°, TR = 10 ms, and TE = 2.67 ms.

The primary objective of the scan parameter optimization was to minimize the standard deviation (STD) of the T₂ estimates while maintaining low bias across the simulated range of tissue properties. The explored parameter space included multiple RF phase increments (φ_inc_) for each steady-state and various odd-to-even flip angle ratios.

The selected parameter set for the two dual-steady state scans consisted of α_odd_ = 12°, α_even_ = 24° with φ_inc_ = 3° for the first dual steady-state and α_odd_ = 19°, α_even_ = 38° with φ_inc_ = 0.5° for the second. This configuration was found to provide a favorable trade-off between precision and bias. A 5-second dummy scan was included in all acquisitions.

### Phantom Imaging

All imaging in this study was performed on a Siemens 7T MAGNETOM Terra MRI system (Siemens Healthcare, Erlangen, Germany). Phantom experiments included a multi-compartment tubes phantom providing a range of T_2_ and T_1_ values, as well as a 3D head-shaped phantom designed to mimic the B_1_ field distribution of the human head while maintaining uniform T_2_ and T_1_ properties.

#### Tubes Phantom

A multi-compartment tubes phantom consisting of four T_2_ and three T_1_ compartments was prepared using varying concentrations of agar and gadolinium. The TWISTARE scan parameters were axial orientation, field of view (FOV) = 150 × 180 × 150 mm³, resolution = 1×1×2.5 mm³, TR=10 ms, TE=2.62 ms, bandwidth (BW) per pixel= 300 Hz/pixel. Separate scan duration was 2:59 minutes, resulting in a total scan time of 5:58 minutes. As a gold standard, a single-echo spin-echo (SE-SE) sequence was acquired in the same axial orientation with FOV = 149 × 149 × 150 mm³ and the same spatial resolution. Additional SE-SE scan parameters were TR= 6500 ms, 3 scans with TE=10,30,50 ms, x3 in-plane acceleration, total scan duration of 20:27 min. To reduce ringing artifacts arising from the tube boundaries, a Hanning filter was applied to the TWISTARE data.

#### Head-Shaped Phantom

A 3D head-shaped phantom was used to mimic similar T_2_, T_1_ and B_1_ field distribution to the brain^51^. Scans were acquired using both positive and negative phase increment configurations, resulting in three separate estimation approaches: positive-ϕ_inc_, negative-ϕ_inc_, and combined. Scan parameters were: sagittal orientation, FOV = 220 × 220 × 168 mm³, isotropic resolution = 1.5 mm³, TR=10ms, TE=2.51 and 6 ms, BW per pixel =300 Hz/pixel. Separate scan duration was 5:22 minutes, resulting in a scan time of 10:44 minutes for positive-ϕ_inc_ and negative-ϕ_inc_, and 21:28 minutes for the combined estimation. The SE-SE scan parameters were sagittal orientation, FOV = 220×220×168 mm^3^, resolution = 1.5 mm isotropic, TR=6500 ms, TE=10,30,50 ms, x3 in-plane acceleration, with a total acquisition time of 20:27 minutes. In addition, a vendor-provided RF field mapping sequence was acquired to compare the resulting RF map with that obtained using TWISTARE. The scan parameters of this scan were FOV = 220×220×168 mm^3^, resolution = 2.3×2.3×4 mm^3^.

### In Vivo Scanning

Human scanning of nine volunteers (4 males, 5 females, aged 22–40 years) was acquired. This study was approved by the Internal Review Board of the Wolfson Medical Center (Holon, Israel) and all scans were performed after obtaining informed suitable written consents. Scans were conducted using the Siemens 7T MAGNETOM Terra MRI system, employing the same scan parameters used in the head-shaped phantom studies: sagittal orientation, FOV = 220×220×168 mm³, and isotropic resolution = 1.5 mm. In two subjects the flip angles of the scan with φ_inc_ = 0.5° were lowered to α_odd_=16°, α_even_=32° due to SAR limitations. These subjects were excluded from per region evaluations. The total scan time for the TWISTARE sequence was 10:44 minutes for a set of two scans. Scans with positive and negative phase increments were collected, this combined set duration was 21:28 minutes. For comparison, a SE-SE reference sequence was also acquired (with the same scan parameters as in the head-shaped phantom set). The vendor RF field map scan parameters were the same as in the head shaped phantom, FOV = 220×20×168 mm^3^ and resolution = 2.3×2.3×4 mm.

#### Estimation of T2 values per brain region

To further assess the performance of TWISTARE, regional T_2_ values were extracted from 94 cerebral regions defined by the Automated Anatomical Labeling (AAL3) atlas^52^. Image preprocessing, including co-registration and atlas normalization, was performed using SPM12^53^. 75 regions containing at least 100 voxels were included in the comparative analysis between methods.

## Results

### 1. Simulation Results

Simulated precision and bias maps of the estimated T_2_ and α for range of values were examined as function of T_2_ and α. Across the range of values relevant to the brain (T_2_ ∈ [15–60 ms and α ∈ [7°–15°]), the bias in T_2_ estimation remained below 4%, and the coefficient of variation was less than 5% (see Supplementary Information Fig. S1). For representative brain tissue parameters (T₂ = 40 ms, α = 10°), the estimated T_2_ value was 40 ± 0.55 ms, and the estimated α was 10 ± 0.11°, showing no bias and <1.5% precision for both T_2_ and α.

### 2. Phantom Imaging – Tubes phantom

A set of tubes with nine components was used to evaluate the experimental precision and bias of TWISTARE. For each component, a region of interest (ROI) was selected to extract mean R_2_ (R_2_=1/T_2_) values and their standard deviations. A linear fit between agarose concentration and the average R_2_ per component was added (Fig.3). A bias of 2 [s^-1^] (<10% from average R_2_) between the linear fit for R_2_ with TWISTARE compared to SE-SE was measured. The ratio between slopes of the TWISTARE and SE-SE linear fits remained near unity (0.994), indicating that the relative scaling of the estimates was preserved. The coefficient of variation across 8 ROIs (excluding ROI #9) was 1.197±0.315% for TWISTARE and 2.130±0.481% for SE-SE, while the TWISTARE acquisition was 3.43-fold faster than SE-SE. This improvement is particularly evident in the central regions (ROI #8 and ROI #9) of the phantom, which contained 3% agar. These ROIs represent areas with pronounced B_1_ inhomogeneity. In these regions, TWISTARE was minimally affected, while SE-SE exhibited significant bias in R_2_ estimation (5.67 s^-1^) and substantially higher variability, with the standard deviation increased by 2.3-fold in ROI #8 and 2.9-fold in ROI #9. The R_2_ values from these ROIs were therefore excluded from the SE-SE–based linear fit to avoid biasing the comparison.

**Figure 3:**
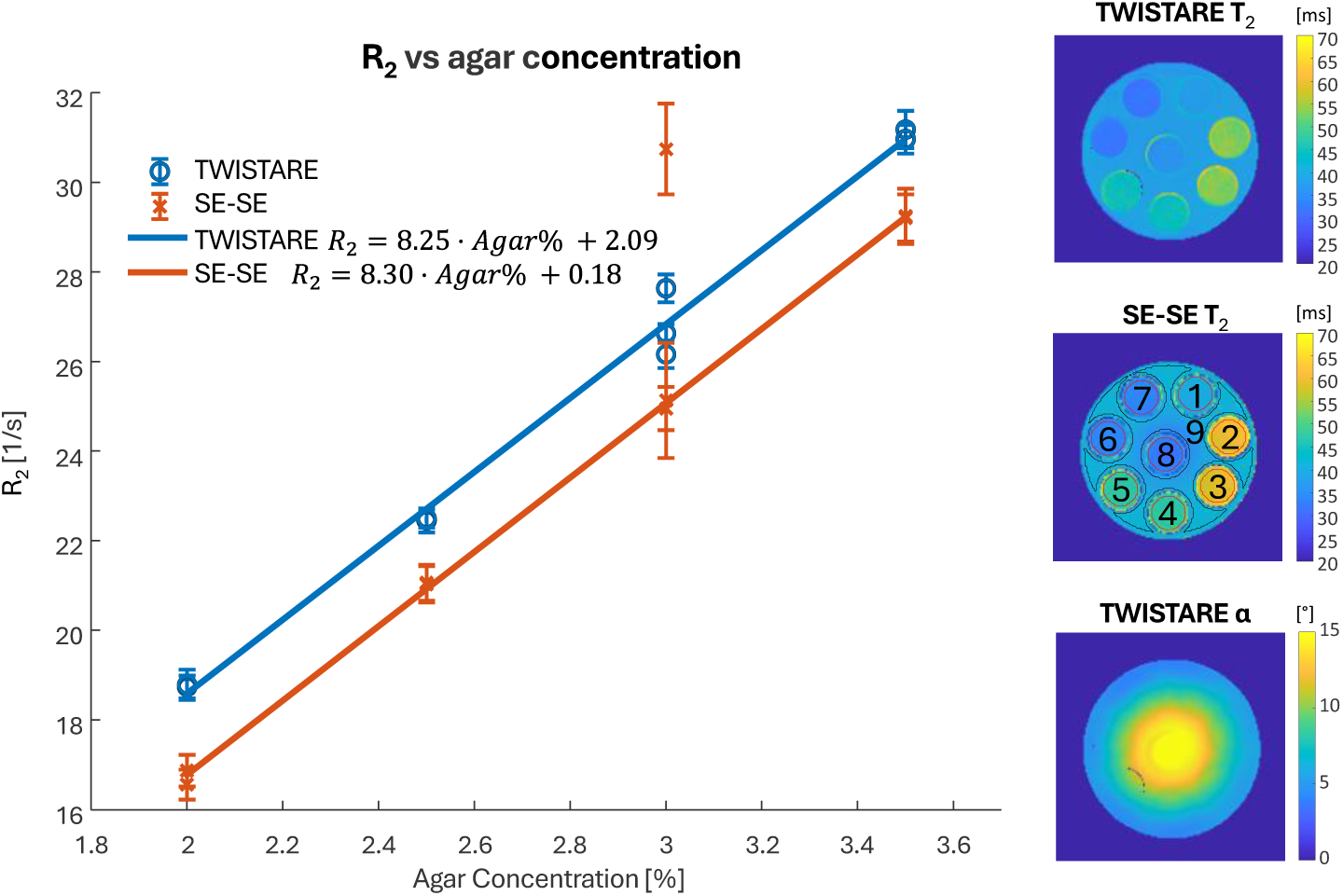
Comparison of T_2_ measurements using TWISTARE and the reference SE-SE method in a multi-compartment tubes phantom. Left- R_2_ (from both TWISTARE and SE-SE) versus agar concentration with linear regression. Right- T_2_ maps from TWISTARE and SE-SE, and the TWISTARE-derived B_1_ map. ROIs corresponding to each compartment are shown on the SE-SE T_2_ map. In regions with strong B₁ inhomogeneity (ROIs #8 and #9), SE-SE showed substantial variability and bias, while TWISTARE was minimally affected.

### 3. Phantom Imaging – 3D head-shaped phantom

A 3D head-shaped phantom with uniform agar-based content — mimicking realistic brain geometry and B_1_ distribution — was used to evaluate the TWISTARE estimation. T_2_ and flip angle maps were generated, and corresponding average values were calculated in the central ROI (see Fig.4) derived with TWISTARE and SE-SE methods. The mean difference in T_2_ values in voxels of the ROI between TWISTARE and SE-SE was <0.67 ms, corresponding to a relative error below 1.72%. The standard deviation of TWISTARE with positive ϕ_inc_ alone was 20% lower than that of SE-SE with half the scan time (Table 1). Combined dataset that included positive and negative phase increment scans resulted in a standard deviation 1.65-fold lower than SE-SE, acquired in a similar time.

**Figure 4:**
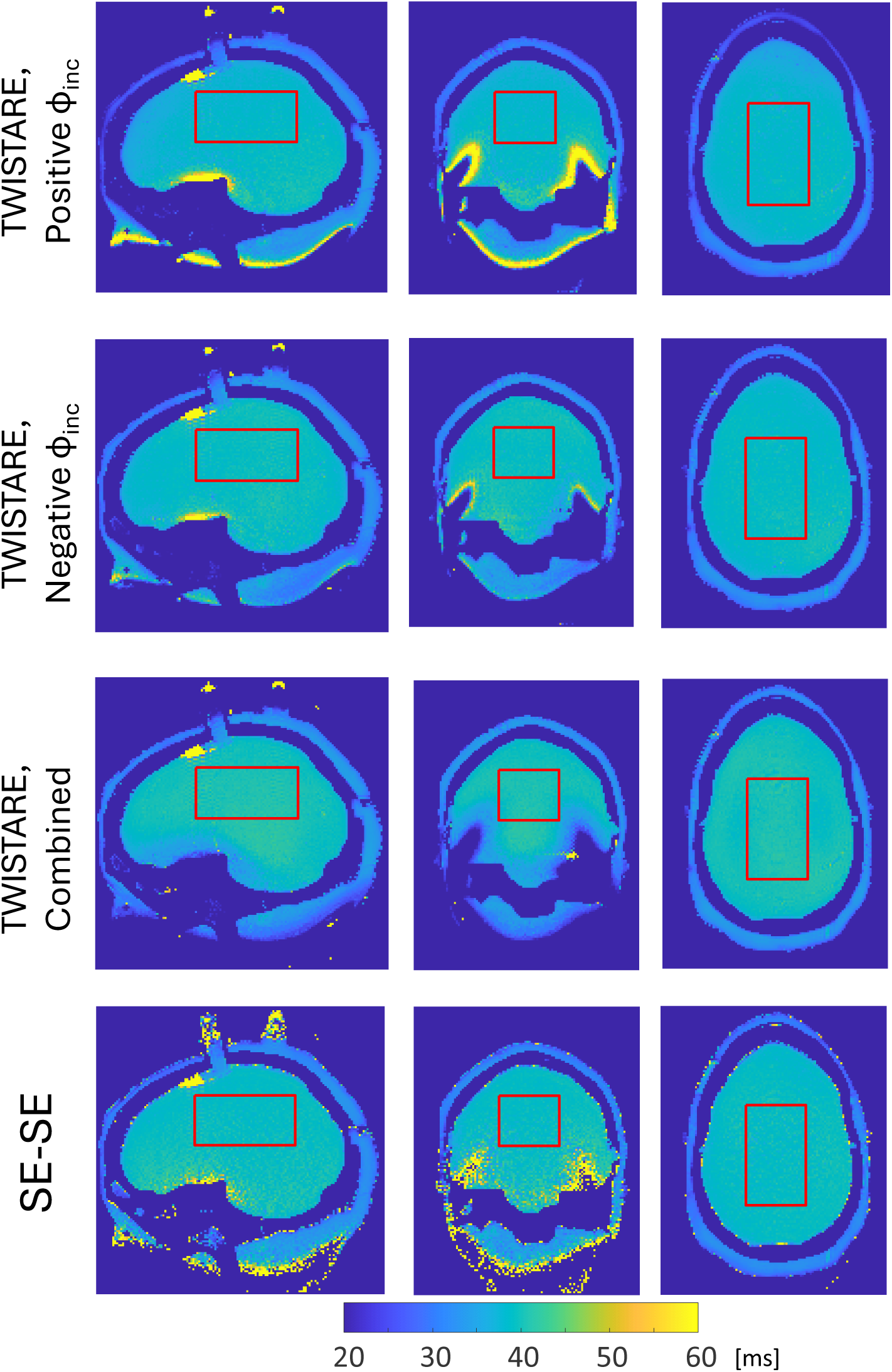
Comparison of T_2_ measurements using TWISTARE and the reference SE-SE method in a 3D head-shaped phantom. From top to bottom: T_2_ maps derived from TWISTARE with positive ϕ_inc_, TWISTARE with negative ϕ_inc_, TWISTARE combined configuration and SE-SE. Each map shows three main cross sections and the ROI for which average values were extracted.

**Table 1.** T2 (mean and standard deviation, STD) for 3D head-shaped phantom.

| **Table 1. T2 (mean and standard deviation, STD) for 3D head-shaped phantom.** |  |  |
| --- | --- | --- |
| method | T2 mean [ms] | T2 STD [ms] |
| TWISTARE, positive $\phi_{inc}$ (10:44 min.) | 38.47 | 0.71 |
| TWISTARE, negative $\phi_{inc}$ (10:44 min.) | 39.37 | 0.65 |
| TWISTARE, combined set (21:28 min.) | 38.92 | 0.52 |
| SE-SE (20:27 min.) | 39.14 | 0.86 |

Fig. 5a shows the B_1_ maps estimated with TWISTARE compared to the vendor-provided RF map. Within the central “brain” region of the head-shaped phantom (Fig.5a), TWISTARE B_1_ estimates obtained with positive ϕ_inc_ alone showed good agreement with the vendor-provided map (Pearson correlation coefficient r= 0.99; root-mean-square error (RMSE) = 0.36°; median absolute percentage error = 4.32%).

**Figure 5:**
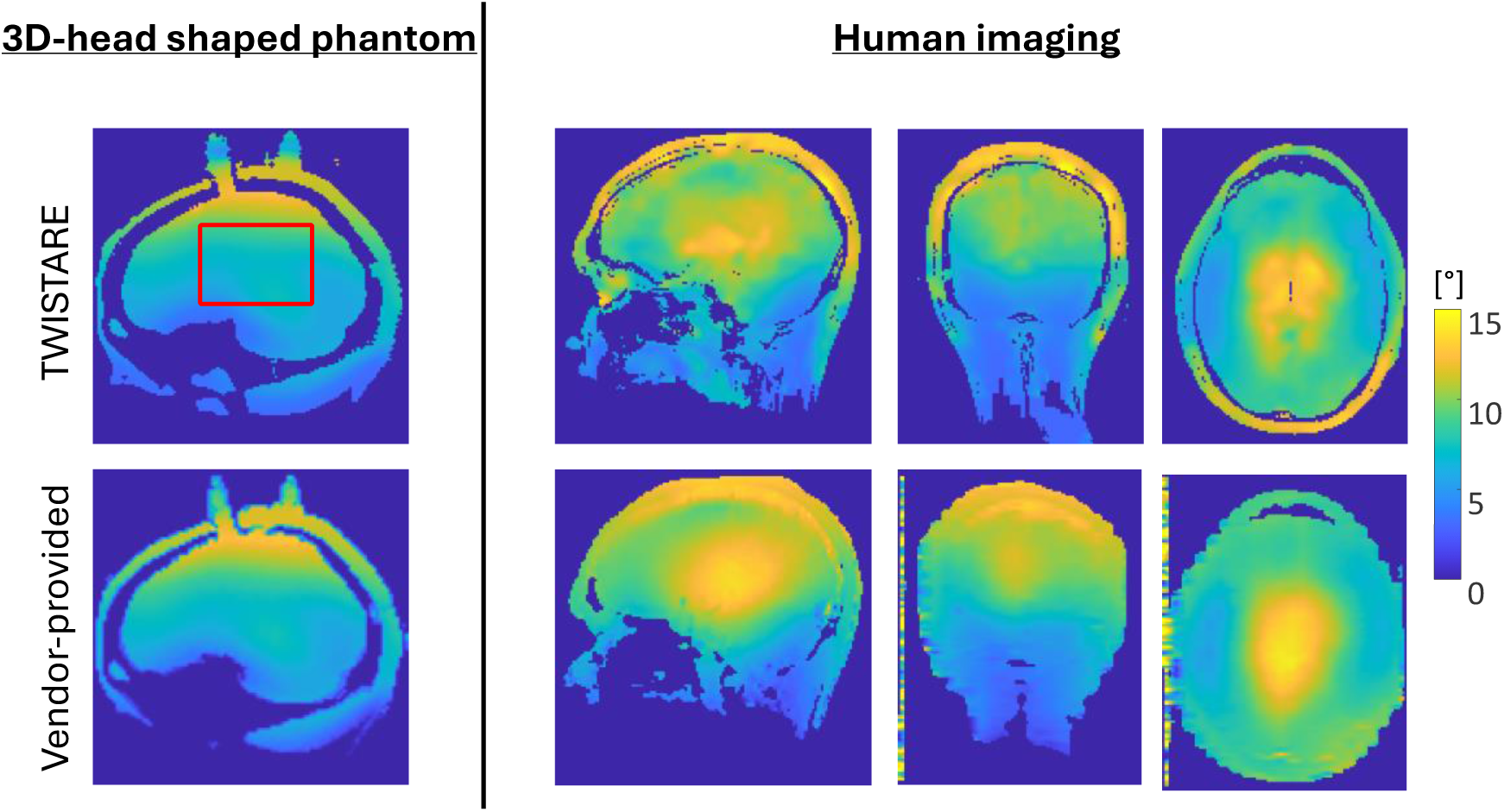
Comparison of B₁ maps estimated with TWISTARE and with the vendor’s sequence. Left: 3D head-shaped phantom (sagittal plane). Right: Representative human subject (three main cross sections).

### 4. In Vivo Results

The TWISTARE method was further evaluated in healthy volunteers to assess its performance in realistic in-vivo conditions. To highlight the importance of B_1_ mapping for reliable T₂ estimation, TWISTARE was also compared to a single-state method, in which the B_1_ distribution cannot be estimated and is assumed to be uniform and fixed to the nominal scan value. Figure 6 shows representative in-vivo T₂ maps estimated from a single-steady-state scan compared to T_2_ map estimated with the TWISTARE method. For the single-state estimation, the first steady-state (ϕ_inc_=3°) was used. Separate scans with positive and negative ϕ_inc_ were used to eliminate the B_0_ contribution. The T_2_ maps with a single-steady-state show pronounced B_1_-related bias, with significant drop of the estimated T_2_ value in the cerebellum and temporal lobes, as expected due to the B_1_ field inhomogeneity in the brain. In contrast, the TWISTARE method effectively accounts for voxel-wise B_1_ variability, producing reliable T₂ maps free from B_1_ trends.

**Figure 6:**
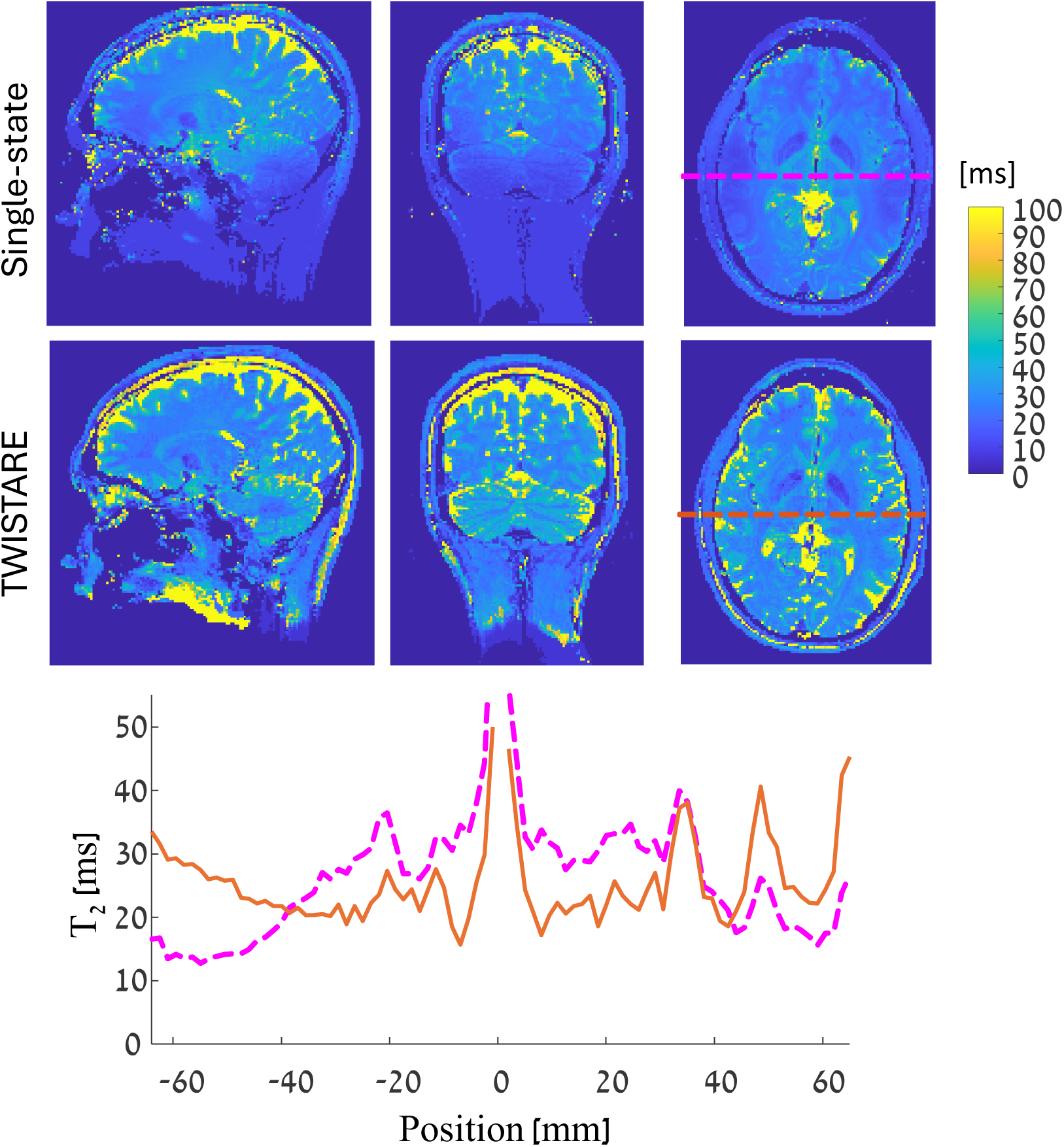
Human imaging - Comparison of T₂ maps obtained using a single-state approach (assuming fixed B₁) and TWISTARE. Three orthogonal cross-sections are shown. For each method, a line profile extracted from the axial slice is displayed, highlighting the T₂ inhomogeneity caused by B₁ variation in the single-state estimation.

Whole-brain T₂ maps with 1.5 mm isotropic resolution were generated using TWISTARE with positive ϕ_inc_, negative ϕ_inc_ and the combined set. The results were compared to the gold-standard SE-SE reference (Fig.7). T_2_ values were analyzed across 75 brain regions in each subject, with each region comprising more than 100 voxels. Median T₂ values estimated with TWISTARE were plotted against those obtained with SE-SE across all 75 regions in four subjects (Fig.8a). A linear fit constrained to zero intercept shows a consistent scaling factor of ∼0.69 between TWISTARE and SE-SE across regions. Histograms of the T_2_ distributions in four representative deep-brain regions - hippocampus, amygdala, putamen and thalamus – are shown in Fig.8b. Table 2 summarizes T_2_ estimates for these regions in one representative subject. The coefficient of variation of T_2_ value was 10-20% for SE-SE and 15-25% for TWISTARE (combined configuration). A consistent deviation was observed between the positive and negative ϕ_inc_ estimates, which may be attributed to residual phase biases that are not fully canceled. Fig. 5b further shows the B_1_ maps estimated with TWISTARE compared with the vendor-provided RF map, demonstrating similar spatial distribution.

**Figure 7:**
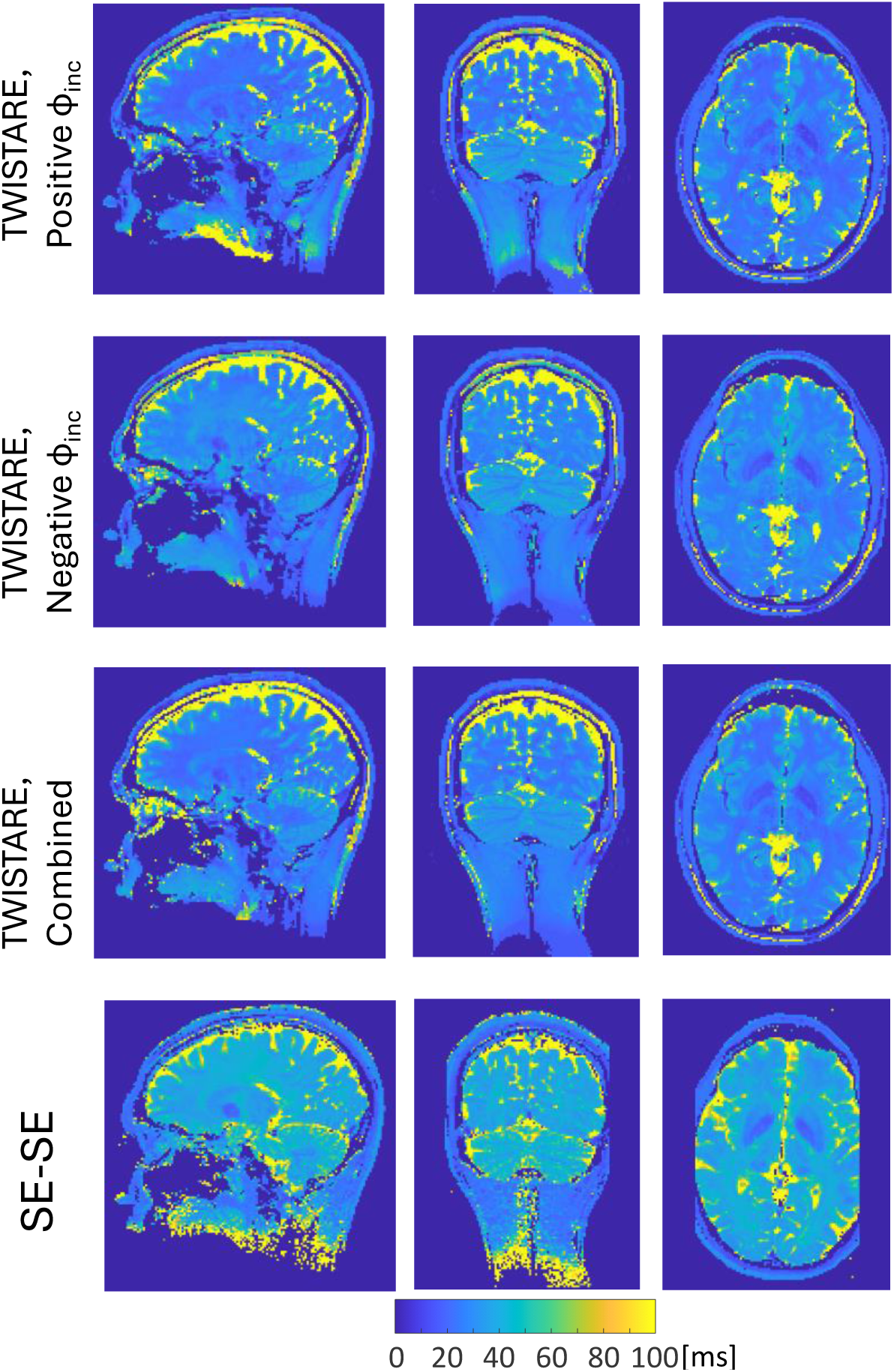
Human imaging - T₂ maps obtained with TWISTARE using three configurations - positive ϕ_inc_, negative ϕ_inc_, and combined - compared with SE-SE. Three main cross sections are displayed.

**Figure 8:**
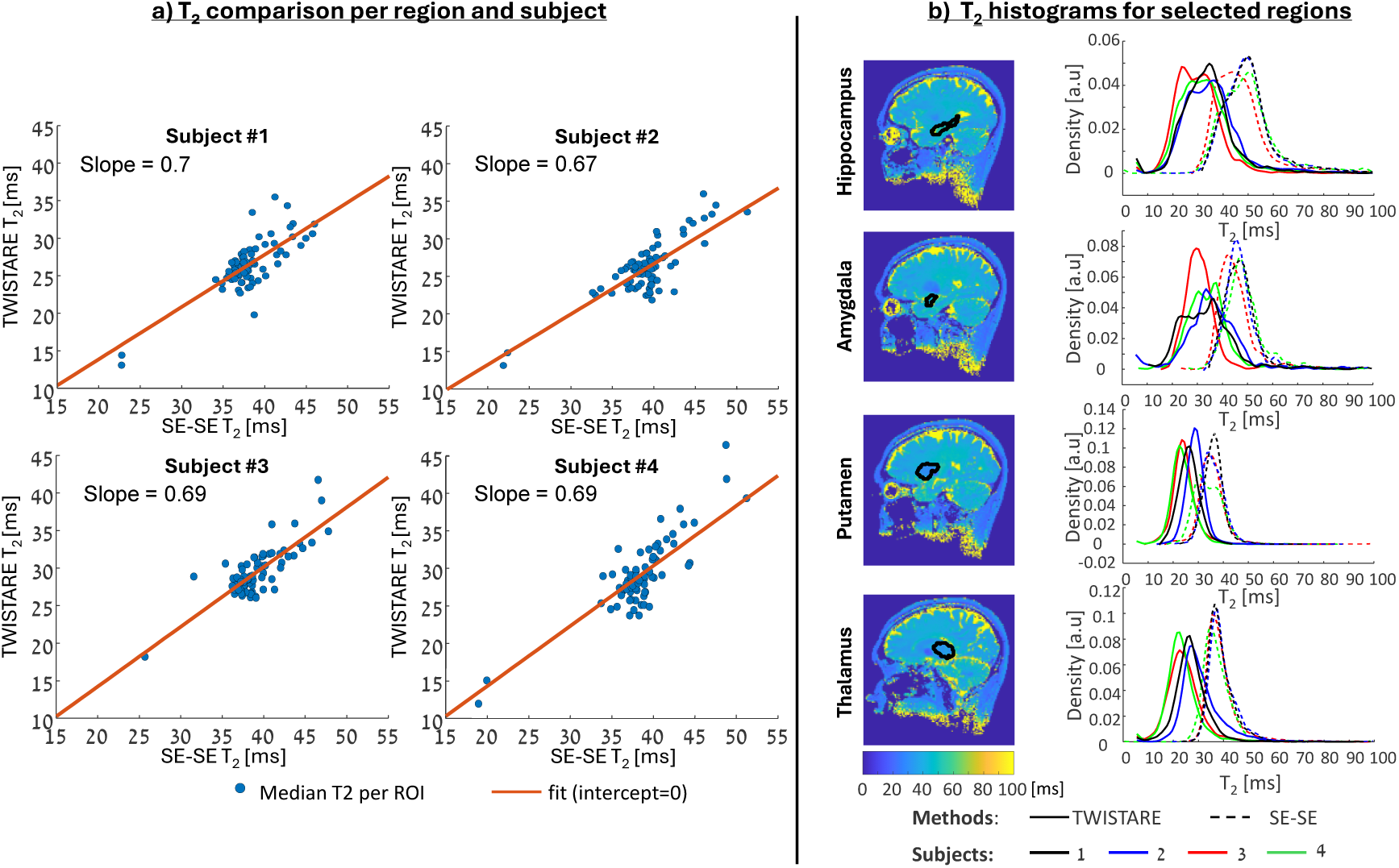
Human imaging - comparison of T_2_ values across brain regions. (a) Median T₂ (TWISTARE vs. SE-SE) across brain regions in four subjects. (b) Representative regions - hippocampus, amygdala, putamen, thalamus - with normalized T₂ distributions (TWISTARE: solid; SE-SE: dashed). Subjects are color-coded.

**Table 2.**
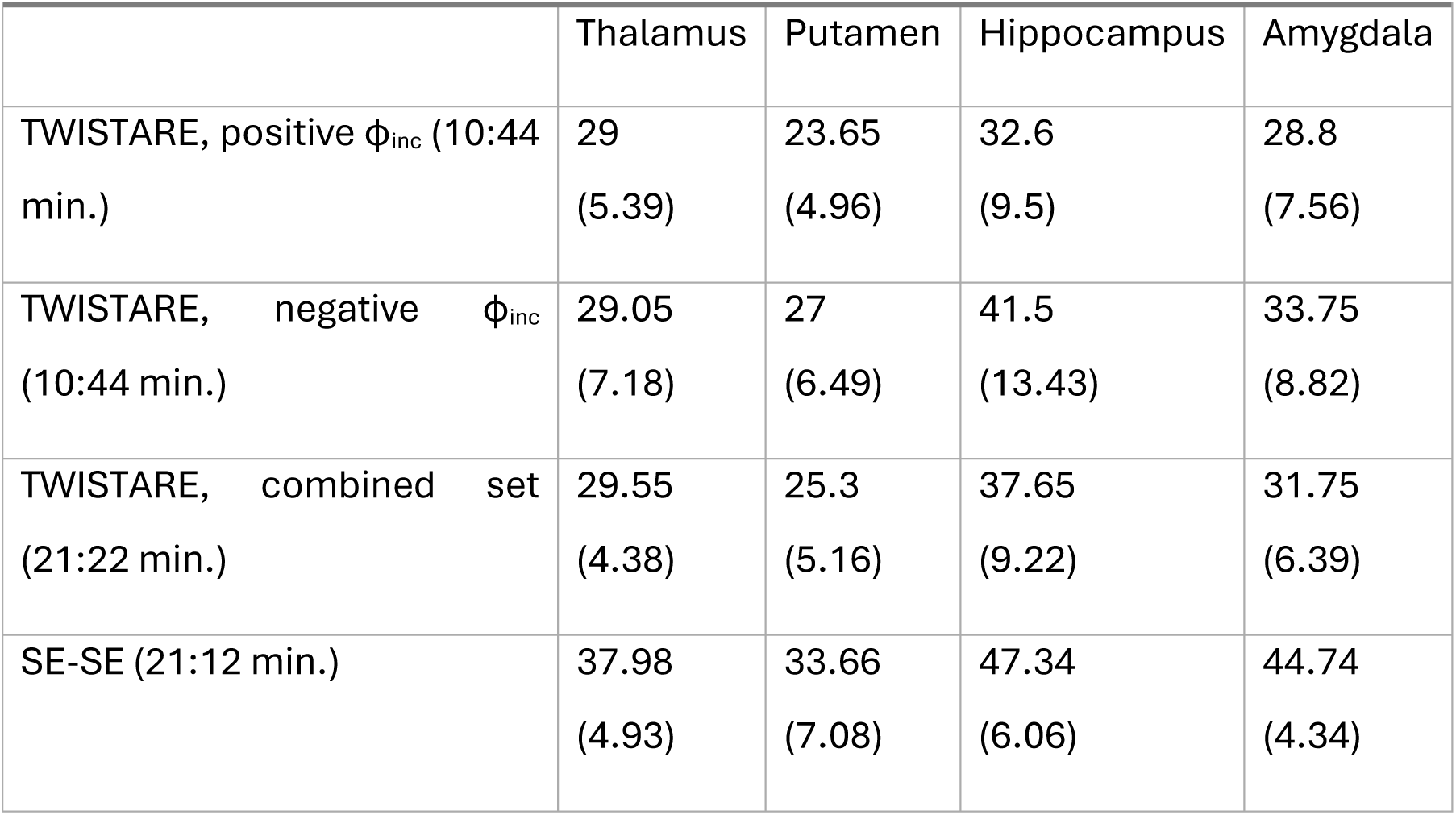
In-vivo T2 estimates [ms] (mean and standard deviation, STD) for four brain regions (estimated in representing subject).

## Discussion

In this study, we introduced TWISTARE, a dual steady-state GRE framework for simultaneous T₂ and B₁ mapping at 7T using only two 3D acquisitions. The proposed method extends phase-based steady-state imaging by interleaving two flip angles. Bloch simulations demonstrated low bias (<4%) and high precision (<5% relative error) across physiologically relevant T₂ and flip angle ranges.

Phantom experiments further validated the method under controlled conditions. In the tubes’ phantom, TWISTARE preserved the relative scaling of R₂ values compared with SE-SE (slope ratio 0.994), while achieving a 3.4-fold reduction in acquisition time. Importantly, in regions with pronounced B₁ heterogeneity, TWISTARE maintained low bias and variance, whereas SE-SE showed elevated bias and variability. This finding highlights the advantage of explicitly estimating flip angle within the reconstruction framework, particularly at 7T where spatially varying transmit fields strongly affect quantitative accuracy.

In the 3D head-shaped phantom, TWISTARE-derived T₂ values agreed closely with SE-SE (<1.72% relative difference), and B₁ estimates showed excellent agreement with the vendor-provided RF map (Pearson r = 0.99). These results confirm that the dual steady-state approach can reliably decouple T₂ and B₁ effects under realistic RF field distributions.

In vivo experiments demonstrated the joint T₂ and B₁ estimation. Single-state reconstructions assuming a fixed flip angle exhibited clear spatial bias, particularly in inferior and temporal regions where B₁ inhomogeneity is most pronounced at 7T. TWISTARE effectively mitigated these trends, yielding spatially consistent T₂ maps across the brain. This confirms that unmodeled B₁ variation remains a dominant source of bias in steady-state T₂ mapping at ultrahigh field.

Regional analysis across in-vivo atlas-defined structures showed strong correspondence between TWISTARE and SE-SE estimates, although a systematic scaling factor (∼0.69) was observed. Several factors may contribute to this discrepancy, including differences in signal modeling assumptions (steady-state GRE vs. spin-echo), sensitivity to stimulated and residual coherence pathways, T₁ insensitivity, magnetization transfer effects, or diffusion-related influences in the SE reference. Importantly, the consistent scaling across regions suggests that TWISTARE preserves relative T_2_ values, which are critical for longitudinal and comparative studies. This deviation is consistent with previous reports using short-TR steady state acquisitions^25,38,41^. A deviation between the positive and negative phase increment datasets was also observed, which may be attributed to residual phase biases that were not entirely eliminated. Future work may explore calibration strategies or refined modeling to further reduce these biases.

Relative to MRF-based frameworks, TWISTARE uses a more constrained acquisition design requiring only two steady-state scans, thereby reducing computational complexity and acquisition time. Although dictionary matching is employed, the parameter space is limited, providing more efficient estimation.

Importantly, TWISTARE improves upon prior phase-based GRE method^41^ that required three to four separate scans to eliminate B₀ contributions. By leveraging intra-scan phase differences between interleaved steady states, the proposed method reduces the acquisition to two scans, improving robustness to motion and B₀ drift while increasing time efficiency.

Future work should extend this framework to enable simultaneous T₁ estimation, which may reduce potential estimation bias and broaden the range of quantitative in vivo biomarkers. Another challenge of the small phase increments is the sensitivity to physiological fluctuations artifacts and undesired stimulated echoes that can manifest. To cope with these challenges acquisition ordering^54^ and flip angle optimization^32^ schemes may help.

In conclusion, TWISTARE provides a time-efficient dual steady-state framework for simultaneous T₂ and B₁ mapping at 7T. By combining interleaved flip angles scans and Bloch simulation modeling, the method provides T₂ estimation while mitigating B₁-related bias. The reduction from four to two acquisitions enhances robustness and practicality for high-resolution 3D brain imaging. TWISTARE offers a new direction using fast steady-state approaches for quantitative neuroimaging at ultrahigh field.

## Acknowledgments

We are grateful to Dr. Amir Seginer for insightful discussions, and to Dr. Sagit Shushan (Wolfson Medical Center), Dr. Edna Haran and the Weizmann Institute’s MRI technician team - E. Tegareh and N. Oshri - for their assistance with the human imaging scans. Dr. R. Schmidt’s lab research was generously supported by Mike and Valeria Rosenbloom Center for Research on Positive Neuroscience and Joyce Eisenberg Keefer and Mel Keefer Career Development Chair for New Scientists.

## Supporting Information

Additional supporting information can be found online in the Supporting Information section at the end of the article.

## Data availability

The scripts and a representative dataset generated during this study will be available at https://doi.org/10.34933/199fc450-7960-43e6-8592-93be108976fa.

## Compliance with Ethical Standards

### Funding

This study was supported by the Mike and Valeria Rosenbloom Center for Research on Positive Neuroscience.

### Ethical approval

All procedures performed in studies involving human participants were in accordance with the ethical standards of the institutional and/or national research committee and with the 1964 Helsinki declaration and its later amendments or comparable ethical standards.

### Informed consent

Informed consent was obtained from all individual participants included in the study.

## Supplementary Information for

### S1. Weights of the estimation algorithm

The inputs to the TWISTARE estimation algorithm are twelve observables generated from the different scans and states, they consist of six phase differences 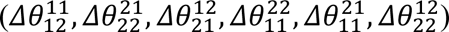 and six magnitude ratios 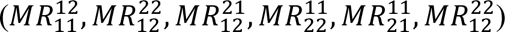. Since these observables (*O_i_*, *i*=1,…,12; where ***O*** denotes the full observable vector) differ both in numerical scale and in their sensitivity to the parameters T_2_, T_1_, and the flip angle (α), a weighted Euclidean distance (d_w_) was introduced. This weighted distance balances the different contributions of the *measured* observables ***O*** during nearest-neighbor matching to observables ***D***((*α*, *T*_2_, *T*_1_)) in the dictionary (also with twelve components *D_i_*, i=1,…,12). The dictionary was generated from Bloch-simulations, determining an observable ***D***((*α*, *T*_2_, *T*_1_)) for every (*α*, *T*_2_, *T*_1_) combination in the parameter space *S*_*α,T*2,*T*1_ covered by the dictionary. The *d_w_* metric with weights *w*_*i*_ is expressed as

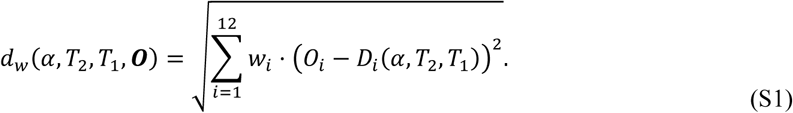

The weights (*w*_*i*_) were derived from the complex signals, found by Bloch simulations for the dictionary used, after supplementing them with additive Gaussian noise. To ensure statistical robustness, for every observable ***D***((*α*, *T*_2_, *T*_1_)) in the dictionary ((*α*, *T*_2_, *T*_1_)∈*S*_*α,T*2,*T*1_), N=10,000 noise samples were drawn and a set *S*_*n*_ of N measurement-like observables ***O*** was realized. The noise level was selected to correspond to an SNR of 100 in a representative central white matter voxel (*T*_1_ = 1.2 s, *T*_2_ = 38ms), assuming a practical flip angle of 10°, repetition time (TR) of 10 ms, and echo time (TE) of 2.67 ms. Because some observable distributions are highly skewed, the interquartile range (IQR) was used to measure variability, and the median was used for central tendency. Under these conditions, the variability of each observable component *O_i_* was quantified in two different dimensions: across *S*_*n*_ (all realizations) and across *S*_*α,T*2,*T*1_ (all dictionary parameter combinations). The corresponding weights were computed according to Equation S2.

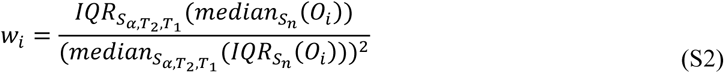

This formulation ensures that observables exhibiting higher stability (corresponding to low IQR over noise realizations) and higher sensitivity (large IQR over the *S*_*α,T*2,*T*1_ space) receive stronger weighting. The proposed weighting scheme appeared to be particularly useful for tissue separation, especially between brain tissue and CSF.

The weights in the metric for the dictionary used in TWISTARE, derived from a two-scan dual-steady state dictionary (Scan 1: α_odd_ = 12°, α_even_ = 24° and φ_inc_ = 3°; Scan 2: α_odd_ = 19°, α_even_ = 38° and φ_inc_ = 0.5°), are provided in Table S1.

**Table S1.**
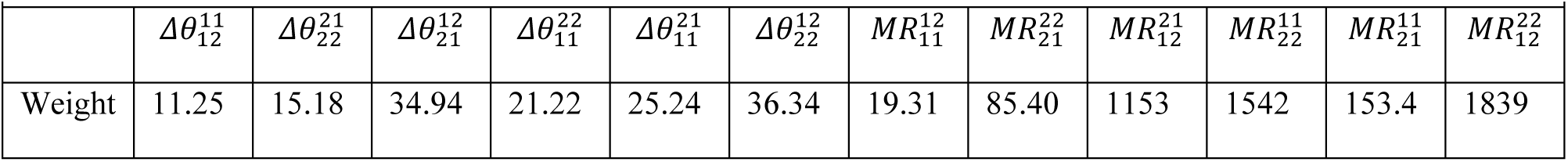
Weights of the observables’ distance metric used in TWISTARE, derived from a two-scan dual-steady state dictionary (Scan 1: α_odd_ = 12°, α_even_ = 24° and φ_inc_ = 3°; Scan 2: α_odd_ = 19°, α_even_ = 38° and φ_inc_ = 0.5°).

### S2. Numerical simulations of the precision and bias of the T_2_ and α estimation

Precision and bias maps of the T_2_ estimations and of the α estimations were derived as function of both T_2_ and α, with T₁ fixed at 1.2 s. Figures S1 shows the resulting maps, including a highlighted window corresponding to physiologically relevant brain tissue values (T₂ ∈ [15–60 ms and α ∈ [7°–15°]).

**Figure S1.**
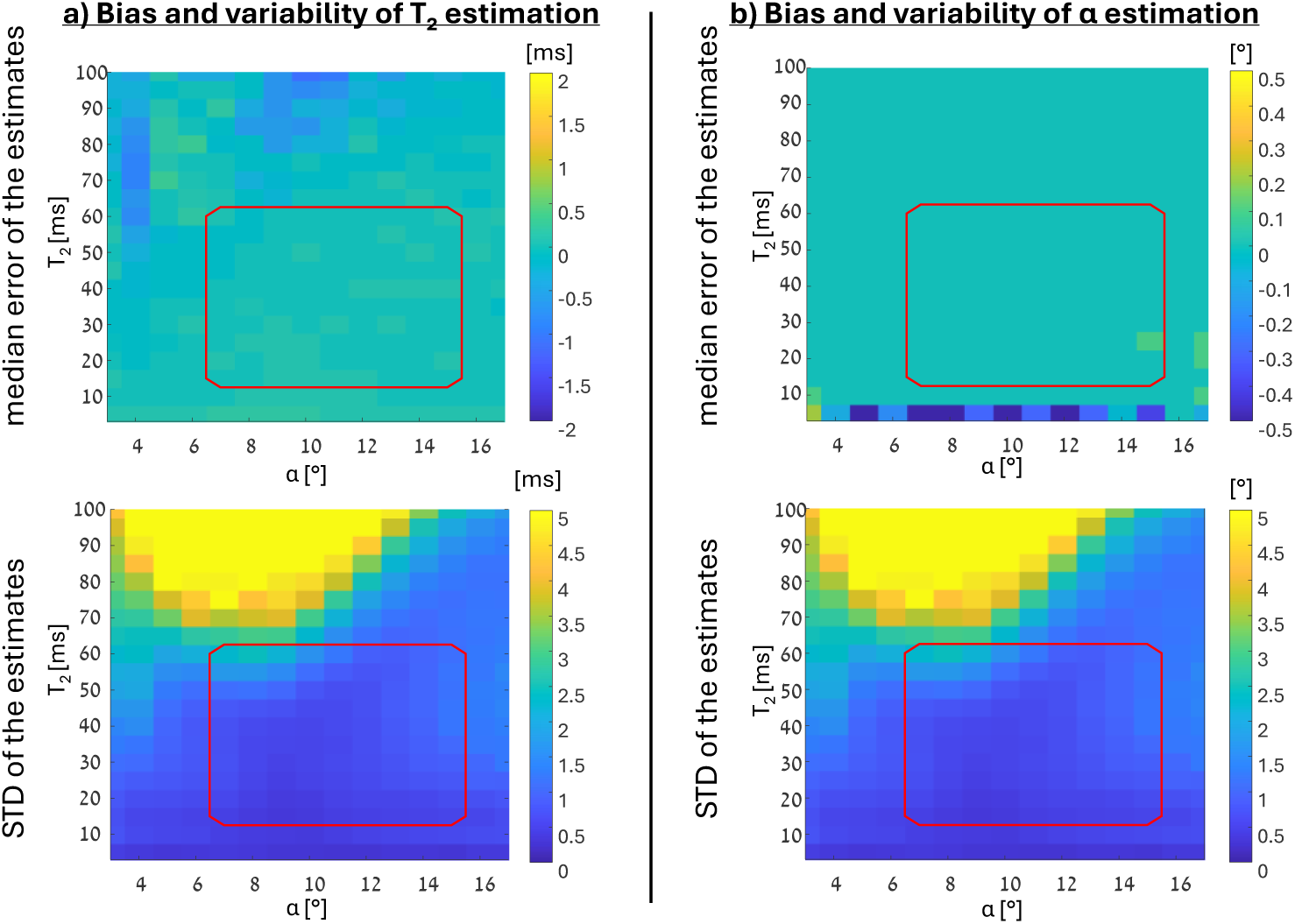
Bias (median error) and variability (standard deviation) of T₂ and α estimates. The red overlay highlights the parameter window corresponding to physiologically relevant values for brain imaging with the proposed method (α 7°–15°, T2 15–60 ms).

